# Subtyping Schizophrenia Using Brain Imaging: A Critical Appraisal of Clustering-Based Models

**DOI:** 10.1101/2025.08.22.25334263

**Authors:** Yuetong Yu, Ruiyang Ge, Sophia Frangou

## Abstract

**Background:** Efforts to define biologically grounded subtypes of schizophrenia have increasingly leveraged neuroimaging data and clustering algorithms. Such approaches aim to capture patient-level heterogeneity with potential clinical and mechanistic relevance. This review evaluates whether structural neuroimaging-derived subtypes can be robustly identified and meaningfully linked to clinical variation.

**Methods:** A systematic review was conducted of peer-reviewed studies published between January 2015 and December 2024 that applied data-driven clustering algorithms to neuroimaging data to identify patient-level subtypes of individuals with schizophrenia or related spectrum disorders. Transdiagnostic studies and those focusing solely on case-control classification, or on feature-level clustering without individual-level subtype assignment, were excluded.

**Results:** Eighteen studies met inclusion criteria. Most used structural MRI, but input features and clustering algorithms varied widely. Across studies, three broad neuroanatomical patterns were described: subtypes with widespread reductions in brain structure, those with regionally circumscribed abnormalities, and those with largely preserved profiles. However, the specific brain regions implicated within each category varied considerably between studies, and no subtype profile was consistently reproduced. Subtypes were not reliably associated with clinical features although there was a trend for higher clinical burden for the widespread subtypes.

**Conclusions:** Current evidence is insufficient to determine whether macroscale neuroimaging features can define subtypes of schizophrenia that are biologically valid or clinically meaningful. Given the limited and inconsistent findings, the subtypes reported to date may reflect continuous variation within the disorder rather than discrete, biologically distinct entities. Advancing the field will require larger, harmonized datasets, standardized analytic pipelines, and rigorous external and longitudinal validation.

## Introduction

Biological subtyping seeks to replace symptom-based taxonomies with classifications grounded in measurable biological features. There are broadly two approaches to subtyping: one that identifies patterns of heterogeneity across individuals, offering insights into group-level variation but lacks direct applicability to individuals; and another that defines subtypes as clinically meaningful categories assignable at the individual level, with the potential to inform prognosis and support personalized or stratified care. The latter approach that focuses on biologically informed individual-level categorization is central to the paradigm of precision medicine. Cancer care leads the field, with molecular subtypes already embedded in diagnostic and treatment protocols (1,2). A comparable emphasis on individual-level categorization dominates recent clustering efforts in neurology. In conditions such as multiple sclerosis (3) and Parkinson’s disease (4), accumulating evidence supports the existence of biologically distinct subgroups of patients identifiable through neuroimaging and computational modeling. This individual-level orientation is fundamental to the logic of clinical practice and underlies the success of biological classifications used in medicine.

Among psychiatric disorders, schizophrenia has long been regarded as a prototypically heterogeneous syndrome, characterized by inter-individual variability in positive and negative symptoms, cognitive deficits, and longitudinal course (5,6). Neuroimaging has played a pivotal role in establishing schizophrenia as a brain disorder by revealing reproducible alterations in brain structure, function, and connectivity (7,8,9). Neuroimaging has also been the most extensively used modality in efforts to identify biological subtypes of schizophrenia (10) largely because its measures are considered relatively stable and more proximally related to underlying neurobiological mechanisms than clinical or behavioral assessments. The increasing availability of large-scale neuroimaging datasets, coupled with advances in standardized preprocessing pipelines, has further established neuroimaging as a central data source for biological subtyping. A wide range of clustering approaches have been applied to neuroimaging data in an effort to delineate subtypes of individuals with schizophrenia based on shared brain-based characteristics that may support clinical stratification or point toward distinct pathophysiological or etiological mechanisms within the broader diagnosis.

This review synthesizes findings from clustering studies in schizophrenia that use neuroimaging features. We focus on clustering approaches that delineate brain-based patient subgroups, rather than latent structures among neuroimaging features, as this distinction is central to the clinical applicability of subtyping frameworks. We review studies published between January 1, 2015, and December 31, 2024 to capture the most recent and methodologically advanced applications and evaluate whether neuroimaging-derived subtypes can be robustly identified and meaningfully linked to clinical variation within schizophrenia. Our appraisal highlights methodological strengths, limitations, and reproducibility issues to inform future research on neuroimaging grounded patient stratification.

## Methods

This review evaluated original studies peer-reviewed published in English in the last ten years (January 1, 2015 to December 31, 2024) and referenced in the major databases (details in supplementary material). We selected: (1) studies that applied clustering algorithms to group individuals with schizophrenia and spectrum disorders into discrete or probabilistic subtypes; studies employing clustering to identify population-level patterns of neuroimaging features without deriving participant-level subtype assignments were excluded; (2) studies that used only neuroimaging features as input to the clustering algorithm were included; studies that incorporated additional modalities (e.g., clinical, cognitive, or genetic data) as input features to the clustering algorithms were excluded. This restriction was imposed to allow an evaluation of the capacity of neuroimaging features to capture biologically meaningful heterogeneity within schizophrenia; multidomain input features would have made it difficult to isolate the specific contribution of neuroimaging to subtype differentiation; (3) studies that explicitly reported the number and defining characteristics of the resulting subtypes; (4) studies that focused exclusively on classification (i.e., distinguishing individuals with schizophrenia from healthy controls) without deriving within-group subtypes were excluded; (5) studies that applied clustering to mixed samples comprising patients with schizophrenia and either healthy controls or unaffected relatives were excluded, as such designs risk deriving clusters that primarily reflect diagnostic group differences rather than meaningful heterogeneity within the patient population; (6) studies that applied clustering to mixed clinical samples comprising patients with schizophrenia alongside those with other psychiatric diagnoses (e.g., mood disorders) were excluded. This decision was made to avoid conflating transdiagnostic variation with within-diagnosis heterogeneity. In mixed-diagnosis samples, clustering algorithms are likely to capture differences driven by diagnostic differences or similarities, thereby obscuring the disorder-specific neuroanatomical patterns necessary for identifying meaningful subtypes within schizophrenia. This problem is exacerbated when diagnostic groups in mixed samples differ in age, sex, sample size, illness stage, or medication exposure, as these imbalances can bias clustering toward features linked to such confounders rather than to the intrinsic heterogeneity of schizophrenia; (7) preprints, reviews, meta-analyses, commentaries, and letters were also excluded. Titles and abstracts were screened independently by two of the authors to determine eligibility, followed by full-text review of all potentially relevant articles. Disagreements were resolved through discussion and consensus.

## Results

We identified 18 studies that met the selection criteria. The PRISMA Checklist and Flowchart are included as supplementary material. Details of the individual studies and their main findings are presented in Table 1. Below we provide a synthesis of the key observations and findings organized thematically.

### Samples

The studies identified examined a broad range of populations with regards to sample size, demographic and clinical characteristics and geographic origin (Table 1). The size of the patients’ samples across studies was small to modest (range 71-1124). The study by Jiang *et al*. (11) is a notable outlier as it included more than 4,000 patients from 24 independent sites. Some studies focused specifically on first episode or early-stage patients (12,13,14,15) while the majority included established cases or mixed samples. Generally, the samples involved young to early middle-aged adults, with the average ages across studies clustered between 25 and 35 years. The percentage of male participants across samples varied from approximately 49% (15) to 74% (16), with most studies reporting a male predominance (modal range across studies was 60–70% male) (Table 1).

The clinical stage of patients was explicitly defined in the studies that focused on first episode psychosis (FEP) (14,15,17,18). The remaining studies included cases that could be characterized either as early stage (defined here as having average illness duration of less than five years) or established (defined here as having longer illness duration) or a mixture of both. Most studies included patients with varied antipsychotic exposure while few focused on drug-naïve patients (12,14,15). Several investigations were conducted in China (12,13,14,15,17) while other studies, relied on multi-site datasets primarily from institutions based in North America and Europe (11,16,18,19,20).

### Clustering Methods

Studies adopted a wide array of clustering methodologies reflecting evolving computational strategies but dominated by unsupervised approaches (Figure 1A). Among the unsupervised approaches, variants of the K-means algorithm were the most common method. Multiple studies used the traditional K-means clustering (12,17,21,22,23) which is a centroid-based unsupervised algorithm that partitions individuals into k clusters by minimizing within-cluster variance. Because K-means assumes spherical clusters in Euclidean space, it was frequently preceded by dimensionality reduction to reduce noise and facilitate cluster separation, typically using principal component analysis (PCA). Alternative sparse K-means was used to select the most relevant subset of features (22). Another version used was fuzzy c-means clustering (16) that models the uncertainty inherent in neuroanatomical data by assigning degrees of membership to multiple clusters based on proximity to centroids. Other studies (12,14) employed agglomerative hierarchical clustering, an unsupervised algorithm that identifies subtypes by iteratively merging similar data points based on a distance metric. This approach builds a tree-like structure in which individuals are grouped into increasingly larger clusters, and subtypes are defined by selecting a specific level at which to partition the hierarchy. Less common unsupervised methods included the use of biclustered independent component analysis (B-ICA) (24) and Peak Density Clustering (PDC), also known as “clustering by fast search and find of density peaks (15). B-ICA simultaneously clusters subjects and brain regions by identifying individuals highly weighted on specific spatial neuroimaging component. However, this approach left a substantial proportion of the patient sample unassigned (24). The PDC defines cluster centers as peaks in the feature space that exhibit both high local density and a large distance from any other point of higher density (25). The remaining points are assigned to the same cluster as their nearest neighbor of higher density, producing flexible cluster boundaries that do not assume specific geometric shapes. The Subtype and Stage Inference (SuStaIn) is the most recently developed unsupervised algorithm (26) and was applied in the two largest sample studies (11,20). SuStain defines categorical subtypes and also infers disease progression from cross-sectional data using event-based models; these combine pattern recognition with a data likelihood function to estimate the most probable sequence of brain changes, assuming abnormalities progress monotonically in severity. Each subtype is defined by its own sequence of regional changes, and patients are probabilistically assigned to a position along that trajectory based on their imaging profile. Each patient’s imaging profile is defined by deviation scores for each neuroimaging feature relative to a healthy reference group, quantifying divergence from normative expectations. SuStaIn can model only a limited number of variables, typically fewer than 20, because higher-dimensional input exponentially increases possible progression permutations, making efficient exploration computationally infeasible.

**Figure 1.**
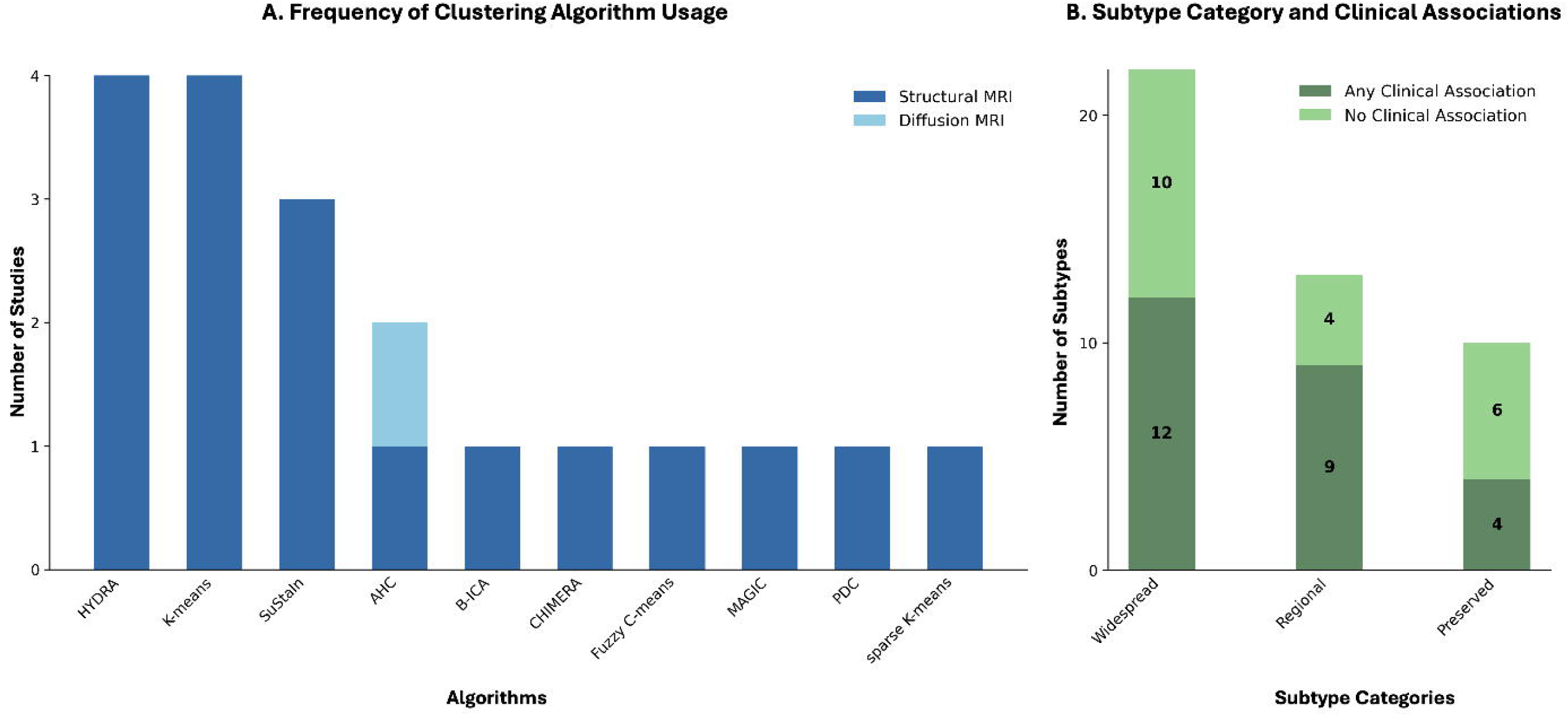
**(A)** Frequency of clustering algorithm use in neuroimaging studies, separated by MRI modalities. **(B)** Number of neuroimaging-derived subtypes (Widespread, Regional, Preserved) according to the presence or absence of clinical associations.

Among the semi-supervised options, HYDRA (Heterogeneity through Discriminative Analysis) (27) was the most common choice (13,18,19,28). HYDRA produces probabilistic cluster assignments by optimizing patient–control separation using multiple support vector machine (SVM) classifiers that leveraged control data to isolate disease-related heterogeneity. Another semi-supervised method used was MAGIC (Multi-scale semi-supervised clustering) (29) which is conceptually related to HYDRA but differs in both feature representation and clustering design. While HYDRA applies a max-margin framework directly to imaging features, MAGIC first reduces voxel-wise data into sparse, interpretable components using orthogonal projective non-negative matrix factorization (opNMF). Another study (30) employed CHIMERA (Clustering of Heterogeneous Disease Effects via Distribution Matching of Imaging Patterns) (31) which is a semi-supervised, registration-based clustering framework that models disease heterogeneity by learning nonlinear transformations from healthy control patterns to disorder related patterns. Rather than clustering subjects directly in feature space, CHIMERA estimates a set of deformation fields—each representing a distinct pattern of anatomical deviation—and assigns patients to the cluster whose learned transformation best reconstructs their neuroimaging profile.

Across studies, the number of clusters was typically selected using internal validation metrics such as silhouette width, Dunn index, or stability analyses, with some studies additionally confirming cluster robustness through leave-one-site-out cross-validation or in a few cases with replication in independent samples (Table 1).

### Neuroimaging Input Features

The studies reviewed primarily relied on sMRI data, except for a single study that used diffusion tensor imaging (DTI) measures (14). Across studies, three main types of features were used. Observed measures included regional gray matter volume or concentration derived from parcellations based on atlases such as MUSE, AAL, Brainnetome, or FreeSurfer segmentations (Table 1). Deviation measures, calculated in several studies (11,15,20,22,32), compared patient-level neuroimaging values to healthy reference distributions to yield individualized deviation scores, which were then entered into clustering algorithms. Component-based measures, used less frequently, involved higher-order anatomical features such as source-based morphometry components (24), structural covariance network features (17), morphometric similarity networks (28), and opNMF components (29). The sole DTI study (14) extracted fractional anisotropy (FA) and mean diffusivity (MD) from 18 major white matter tracts.

### Subtype Number and Neuroimaging Patterns

The subtypes identified differed in the magnitude, direction, and spatial distribution of anatomical alterations, but broadly fell into three categories: widespread—characterized by diffuse structural reductions; regional—marked by circumscribed deviations in specific brain areas; and preserved—showing relatively intact anatomical integrity (Figure 1B and Figure 2).

**Figure 2.**
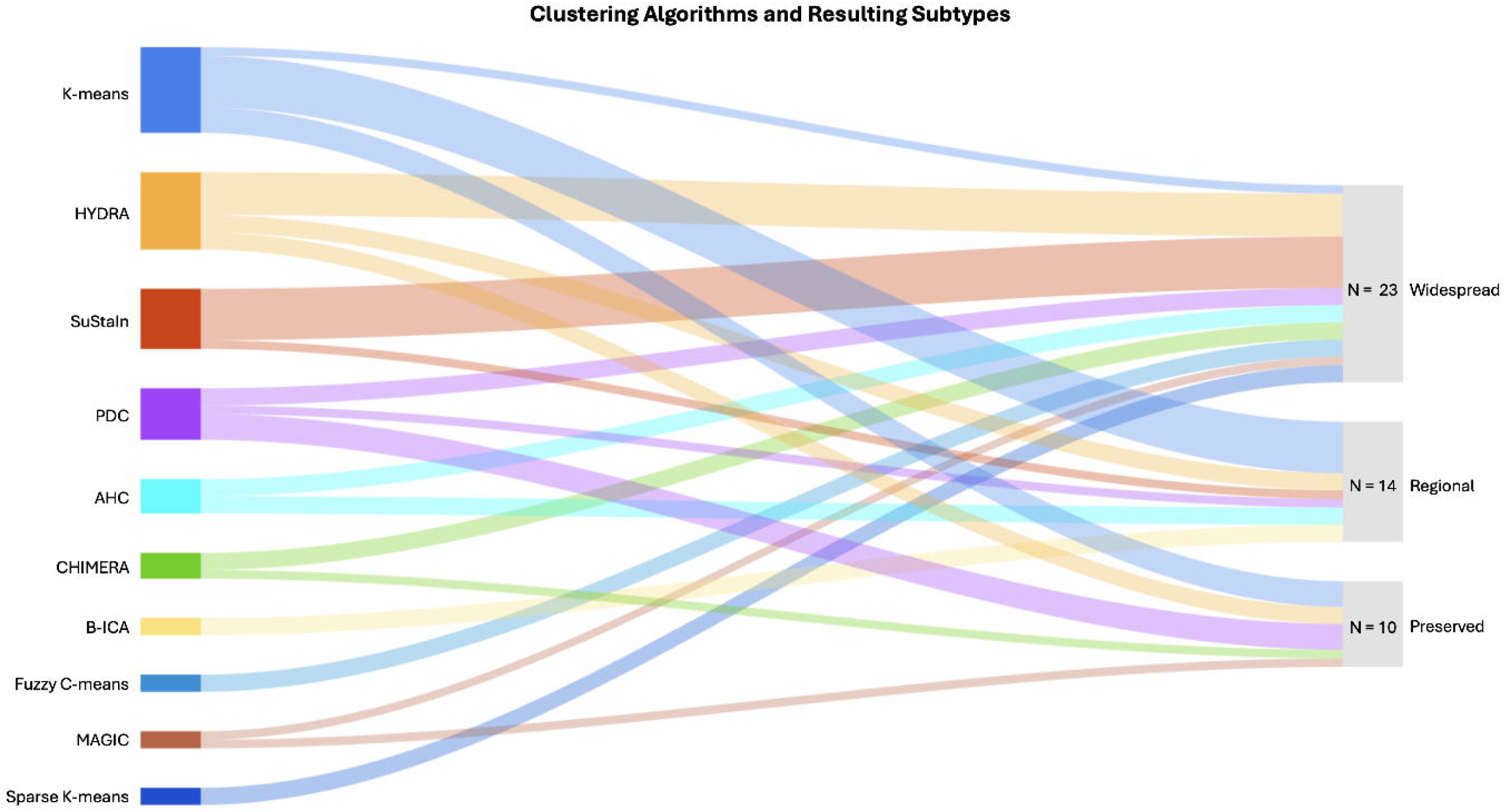
Sankey diagram showing the mapping between clustering algorithms and identified subtypes (Widespread, Regional, Preserved), with flow width proportional to the number of studies. Abbreviations: HYDRA, Heterogeneity through discriminative analysis; SuStaIn, Subtype and Stage Inference; PDC, Peak Density Clustering; AHC, agglomerative hierarchical clustering; CHIMERA, Clustering of heterogeneous disease effects via distribution matching of imaging patterns; B-ICA, biclustered independent component analysis; MAGIC, Multi-scale semi-supervised clustering.

Widespread subtypes were the most frequently reported. These subtypes showed widespread reduction in cortical and subcortical gray matter volume or cortical thickness in patients compared to normative expectations (11,12,13,15,16,18,19,20,29,32) or in morphometric similarity indices (28). Across studies, abnormalities typically encompassed regions within the frontal cortex, superior and middle temporal gyri, thalamus, insula, and hippocampus, although the magnitude and the specific localization of the regional effects varied markedly between studies. Widespread subtypes were observed both in patients with established illness (12,13,15,16,19,29,32) and in first episode/early-stage cases (15,18,28). In the only DTI study (14) the widespread subtype identified demonstrated white matter abnormalities, in FA and MD across major association, commissural, and projection tracts.

Regional subtypes were reported in a smaller subset of studies and were characterized by anatomically restricted deviations in cortical and subcortical regions; the specific regions implicated, and the nature of the alterations showed little consistency across studies or across the clinical stage of the samples involved (11,12,13,14,15,20,21,24,30,32). Examples include subtypes with reduced gray matter concentration in the insula, superior temporal gyrus, and inferior frontal gyrus, or in the superior, middle, and medial frontal gyri (24); reduced gray matter volume in the thalamus, anterior cingulate and superior temporal regions (30); or even more regionally restricted gray matter changes in the supramarginal gyrus, precuneus, and the angular gyrus (21). Other reported patterns included subtypes with selective hippocampal volume loss (15) or volume reductions in other specific subcortical regions (11,20,32), or focal basal ganglia volume enlargement (13) or localized prefrontal and premotor volume increases (12). The only DTI study (14) identified that regional subtype with localized FA reduction in the superior longitudinal fasciculus coupled with increased MD in the corticospinal tract. One study also described a subtype defined by choroid plexus and lateral ventricle enlargement, although its broader anatomical profile was unclear due to limited structural features assessed (23).

A small number of studies reported subtypes with largely preserved neuroanatomical profiles (15,17,18,19). These included patients with established illness (19) or first episode psychosis (18) showing preserved gray and white matter volume. Other studies identified preserved subtypes based on preserved covariance patterns in gray matter volume (15) or structural dissimilarity measures (17), with such profiles observed in both early-stage and established cases.

The SuStaIn-based subtypes identified by Jiang *et al*. (11,20) require further clarification. These studies yielded the two subtypes which appeared to differ primarily in the sequence and spatial emphasis of progressive gray matter loss. Both subtypes began as regionally circumscribed; in one, early changes were concentrated in prefrontal regions before extending to widespread cortical and subcortical areas and in the other, subcortical alterations predominated initially and subsequently progressed to a more widespread pattern. Using the same method in a different sample, Sone *et al*. (32) identified three subtypes defined by initially selective subcortical reductions, widespread cortical thinning, or a combined profile that also included pallidal volume increase.

### Associated Features

Subtype differences were examined in demographic and clinical characteristics and functional outcomes. However, there was substantial inter-study variability in the selection and reporting of these associated features.

Age and sex were evaluated in nearly all studies (Table 1), and the vast majority found no significant differences between subtypes, regardless of whether cohorts comprised early-stage (15,17,18,22,28) or established patients (11,13,14,19,23,29).

Across studies, subtype differences in clinical characteristics were generally absent irrespective of the clinical stage of the samples. Multiple investigations reported no differences in positive or negative symptom scores (11,12,13,15,17,19,21,23,24,28,29,30), illness duration (11,17,19,22,23,29,30), or antipsychotic dose (15,16,19,22,29,30). When differences were present, they indicated that the widespread subtypes had higher symptom scores (16,18,20,22), longer illness duration (13,16) and lower global functioning (14,29). Conversely, preserved subtypes generally showed more favorable clinical characteristics (11,18,20).

## Discussion

This review synthesized recent neuroimaging-based clustering studies aimed at defining biologically grounded subtypes of schizophrenia, focusing on approaches that assign subtypes at the individual level. Across studies, there was substantial variability in sample characteristics, neuroimaging features, and clustering methodologies. Although the specific brain regions implicated varied considerably between studies, three broad conceptual patterns emerged: representing subtypes with either widespread or more regional abnormalities, and subtypes with preserved neuroimaging profiles. Associations with demographic and clinical variables were uncommon, but when present, they tended to link the widespread subtypes to greater clinical severity.

### Methodological Considerations

Several methodological factors warrant consideration when interpreting this subtype literature. Foremost is the adequacy of sample size for reliable subtype identification; most datasets used to date were modest in scale, with only one study including ∼4,000 patients (11). Such limited sample sizes can constrain the statistical power necessary to detect smaller subgroups or those with subtle alterations. This limitation is particularly consequential in the context of schizophrenia, where even case–control neuroimaging differences tend to be modest in magnitude (8,9,33), suggesting that within-diagnosis variation across putative subtypes is likely to be smaller still.

Most studies relied on sMRI as the input modality. While this offers broad availability and well-established pipelines for extracting measures such as cortical thickness, surface area, and subcortical volume, these macroscale features may not fully capture the neurobiological variation most relevant to schizophrenia. The lack of consistency in the neuroimaging profiles of subtypes across studies is likely to reflect the variation in input features. Methodological choices, including the dimensionality reduction techniques, parcellation scheme, and normalization procedures, also varied widely and can substantially influence subtype configurations. These sources of variability were rarely examined systematically. A small number of studies incorporated higher-order sMRI features, such as source-based morphometry components (24), structural covariance dissimilarity matrices (17), morphometric similarity network indices (28), or orthogonal projective non-negative matrix factorization components (29). Although potentially informative, these representations are often harder to interpret anatomically, their selection was seldom justified, and, in this review, they did not yield subtypes that were clearly superior or more clinically informative than those based on conventional region-level measures.

The lack of inter-study consistency in the anatomical configuration of subtypes may also reflect the influence of the clustering methods themselves, as different algorithms and parameter choices can yield markedly different subtype configurations. Most studies used conventional unsupervised approaches such as K-means, hierarchical clustering, or their sparse/fuzzy variants. While computationally efficient, these methods assume simple data geometries (e.g., spherical clusters) and often rely on simple similarity metrics such as Euclidean distance. These assumptions are likely not suited for complex, high-dimensional neuroimaging data, where relationships between individuals are often non-linear or embedded in manifolds that are poorly captured by simple distance-based metrics. As a result, the subtype solutions may be overly influenced by noise or by dominant global features. Semi-supervised approaches like HYDRA and MAGIC, which incorporate healthy control data to isolate disease-relevant variation, introduce different assumptions—such as the separability of patient subtypes from controls using linear or kernel-based decision boundaries—that may not hold in practice. These methods also remain vulnerable to imbalances in confounding variables (e.g., age, sex, scanner type) unless carefully addressed. Furthermore, most algorithms yield hard subtype assignments without membership confidence estimates, limiting the ability to assess reliability or identify unstable classifications.

Without measures that quantify the certainty of subtype membership, it becomes difficult to assess the reliability of group-level comparisons or to identify individuals whose assignment may be unstable or ambiguous.

Given that the two largest studies in this review (11,20) applied the SuStaIn algorithm, its methodological assumptions warrant particular attention. SuStaIn cannot accommodate high-dimensional input because the number of possible progression sequences expands exponentially with each added feature, quickly becoming computationally prohibitive. Consequently, analyses are restricted to a small set of preselected regions. In the studies considered here regions were selected based on case-control differences in univariate analyses which may limit spatial granularity and bias subtype solutions toward the most dominant diagnosis-related effects. SuStaIn relies on normative deviation scores to construct patient-level imaging profiles making results sensitive to the composition of the group used as controls in each study. Study specific recruitment biases of the control group, that are often difficult to detect or quantify, can distort normative deviation estimates leading to spurious abnormalities and, in turn, compromising the validity of subtype assignments. A further consideration pertains to SuStaIn’s inference of longitudinal progression from cross-sectional data, which rests on the assumption that inter-individual variation in patients reflects a fixed, irreversible sequence of accumulating pathology. The algorithm does not account for the alternative possibility that cross-sectional variation in patients’ neuroimaging data may not represent points along a temporal trajectory. Instead, individuals assigned by SuStaIn to more advanced stages may exhibit marked but trait-like deviations that could be present even before illness onset. These factors suggest cautious interpretation of findings using SuStaIn in non-neurodegenerative disorders.

Subtype validation also varied across studies. Most used internal measures such as cluster stability to select the number of subtypes while only a few conducted external validation with independent cohorts (Table 1).

### Clinical Relevance and Interpretability of Neuroimaging-Based Subtypes

Findings from this review suggest that individuals with schizophrenia can be differentiated based on neuroimaging features, yet these distinctions rarely correspond to meaningful differences in clinical presentation. In other words, while clustering consistently detects underlying neuroanatomical heterogeneity, the resulting subtypes do not appear to differ in their clinical presentation. Clinical interpretation is further complicated by the lack of consistent and distinctive anatomical profiles across studies. Even within broad categories such as widespread or regional alterations, the regions involved, and the direction of deviations varied substantially. This anatomical variability limits confidence in the validity of the subtypes identified. Without well-defined and reproducible anatomical profiles, it is challenging to delineate the underlying biological mechanisms of each subtype, and even more difficult to develop them into constructs with clinical applicability.

These observations highlight the challenge of disentangling true biological heterogeneity— reflecting distinct disease mechanisms—from variability along a shared continuum. If true heterogeneity exists, clustering should reveal consistent and orthogonal subtypes with divergent profiles (e.g., one subtype marked by frontotemporal atrophy, another by parieto-occipital changes), each mapping onto distinct clinical signatures. However, the absence of such patterns in this review suggests that the heterogeneity traditionally attributed to schizophrenia based on inter-individual differences in symptom profile, severity, or illness course does not necessarily correspond to distinct underlying brain mechanisms. In this view, much of the observed clinical variability may reflect different manifestations of the same underlying biological processes, that vary in severity influenced by external factors such as developmental stage, environmental exposures, or co-occurring conditions. Alternatively, it may reflect the limitations of macroscale neuroimaging, which yields continuous and collinear features that inherently favor dimensional rather than categorical clustering solutions.

## Conclusion and Future Directions

Current evidence does not yet establish whether macroscale neuroimaging features can define subtypes of schizophrenia that are either clinically or biologically meaningful. While clustering approaches consistently detect underlying neuroanatomical heterogeneity, the anatomical profiles of these subtypes vary substantially across studies, limiting both their interpretability and their potential mechanistic significance. This uncertainty is compounded by persistent methodological challenges, including small and heterogeneous samples, sensitivity to preprocessing and clustering choices, and a lack of external or longitudinal validation. Resolving these issues will require larger, harmonized datasets; rigorous benchmarking of algorithms; and transparent, reproducible analytic pipelines. Determining whether these clusters represent true biological subtypes—as opposed to variation along a shared disease continuum—will also depend on integrating neuroimaging with molecular, genetic, and longitudinal data, and on demonstrating predictive or therapeutic utility.

## Data Availability

All data produced in the present work are contained in the manuscript.

## Disclosures

None of the authors reported any biomedical financial interests or potential conflicts of interest.

## Acknowledgements

None.

## Notes

### Competing Interest Statement

The authors have declared no competing interest.

### Funding Statement

This study did not receive any funding.

