## Supplementary material for "Subtyping Schizophrenia Using Brain Imaging: A Critical Appraisal of Clustering-Based Models": Table 1

| **Table 1. Study Details** | | | | | | | | |
| --- | --- | --- | --- | --- | --- | --- | --- | --- |
| **Study** | **Sample^1^** | **Clinical Stage^2^** | **Input Features** | **Clustering Method** | **Number of Clusters** | **Subtype Description^3^** | **Associated Features ^4^** | **External Validation Sample** |
| **Structural Magnetic Resonance Imaging** | | | | | | | | |
| Gupta et al. 2017^24^ | 382 patients  ~72% male;  ~ Avg Age: 36 yrs | Established | Components of spatial mappings of gray matter concentration derived through ICA from SBM | Biclustered Independent Component Analysis | 2 | Subtype 1: Reduced gray matter concentration in the insula, superior temporal gyrus and inferior frontal gyrus;  Subtype 2: Reduced gray matter concentration in the superior, middle and medial frontal gyri; 202 patients remained unassigned | No subtype differences in age or sex; subtype 2 had higher PANSS positive scores than Subtype 1, with no differences in negative or general scores | No |
| Dwyer et al. 2018^16^ | 145 [71 patients and 74 HC]; ~74% male;  ~ Avg Age: 38 yrs | Established | GMV via VBM | Fuzzy c-means  Clustering after PCA | 2 | Subtype 1: GMV reductions in insula, striatum, thalamus, hippocampus, and right superior temporal regions, with increased volume in medial and lateral parietal lobes;  Subtype 2: GMV reductions in lateral prefrontal, medial parietal, and temporal cortices, with increased cerebellar volume | Subtype 1 had an older age of onset, longer duration of illness and higher PANSS negative symptom score; no differences in antipsychotic dose/type | Yes |
| Honnorat et al. 2019^30^ | 326 [157 patients and 169 HC]; ~ 71% male; ~ Avg Age: 31 yrs | Early Stage | MUSE-derived regional GMV, white matter and CSF volumes | CHIMERA | 3 | Subtype 1: Reduced GMV in the thalamus, anterior cingulate and superior temporal regions and higher CSF volume in temporal regions, WM expansion in temporal regions; Subtype 2: Similar GMV pattern to sutype1 but increase CSF volume in prefrontal regions  Subtype 3: Mild CSF expansion and moderate GM and white matter volume reductions | Subtype 1 had older age and subtype 3 include more females; no subtype differences in illness duration, age of onset, PANSS subscale scores and antipsychotic dose | No |
| Ma et al. 2019^21^ | 67 patients  Unspecified age and sex | FEP drug-naive | GMV of brain regions associated with schizophrenia risk genes | K-means clustering | 3 | Subtype 1: Reduced GMV in the supramarginal gyrus and precuneus and higher GMV in the mid-cingulate cortex;  Subtype 2: Increased GMV in the angular gyrus;  Subtype 3: Subtle GMV reduction of the precuneus | No subtype differences in age and sex; Subtype 1 had higher PANSS positive symptoms scores | No |
| Chand et al. 2020^19^ | 671 [307 patients and 364 HC]; ~60% male; ~ Avg Age: 30 yrs | Established | Regional volume measures of gray and white matter and CSF | HYDRA | 2 | Subtype 1: Widespread volumetric decreases; Subtype 2: Preserved neuroanatomical profile with subtle enlargement of the basal ganglia | No subtype differences age, sex, illness duration, antipsychotic dose/type, age of illness onset, symptom severity | No |
| Liu et al. 2021^17^ | 178 [107 patients and 71 HC]; ~60% male; ~ Avg Age: 25 yrs | FEP drug-naive | IDSCN-derived from covariance matrix of AAL regional GMV | K-means Clustering | 2 | Subtype 1: Preserved covariance profile;  Subtype 2: Decreased covariance between hippocampus/pallidum/putamen and between occipital/orbitofrontal/superior temporal cortical regions | No subtype differences age, sex, illness duration; or PANSS positive/negative symptom scores; the same pattern was identified in 2 FEP datasets; 2 datasets with chronic patients and in a clinical high-risk sample | Yes |
| Chew et al. 2022^13^ | 234 [158 patients and 76 HC]; ~68% male; ~ Avg Age: 32 yrs | Established | FreeSurfer derived regional cortical and subcortical volumes | HYDRA | 2 | Subtype 1: Focal increase in the basal ganglia and third ventricle;  Subtype 2: Widespread cortical and subcortical volume decrease | No subtype differences age, sex, PANSS total, and GAF scores; Subtype 1 longer duration of illness | No |
| Wen et al. 2022^29^ | 1166 (583 patients and 583 HC]; ~62% male; ~ Avg Age: 33 yrs | Established | Components obtained from the T1 images using orthogonal projective non-negative matrix factorization | MAGIC | 2-4 | Plausible solutions identified 2-4 clusters; the 2-cluster solution was chosen as these subtypes were similar to those reported by Chand et al. 2020;  Subtype 1: Widespread gray matter reduction, most pronounced in the insula and thalamus;  Subtype 2: Preserved neuroanatomical profile with subtle enlargement of the basal ganglia | No subtype differences age, sex, PANSS subscale scores, illness duration or age of onset; Subtype 2 had a higher GAF score | No |
| Xiao et al. 2022^15^ | FEP: 336 [163 patients and 173 HC]; ~49% male; ~ Avg Age: 24 yrs  Established: 363 [133 patients and 230 HC]; ~53% male; ~ Avg Age: 33 yrs | FEP drug naïve and established | Dissimilarity measures based on FreeSurfer-derived measures of regional cortical thickness surface area | Density Peak-Based Clustering | 3 | FEP: Subtype 1 showed a widespread decrease in cortical thickness coupled with a focal increase in the rostral anterior cingulate;  Subtypes 2 and 3 had preserved neuroanatomical profiles.  Established: Subtype 1 showed widespread cortical deficits coupled with reduced hippocampal volume and increased pallidal volume;  Subtype 2 had a focal decrease in hippocampal volume; Subtype 3 had a preserved neuroanatomical profile | Within each dataset, subtypes did not differ in age, sex [ except for a slight excess of females in FEP subtype 2], PANSS subscale scores, GAF, and antipsychotic dose | Yes |
| Chai et al. 2023^12^ | 571 [314 patients and 257 HC]; ~69% male; ~ Avg Age: 34 yrs | Mixed: Established and drug naive FEP patients | GMV via VBM | K-means and Agglomerative Hierarchical Clustering | 2 | Subtype 1: Widespread GMV decrease;  Subtype 2: Focally increased GMV in prefrontal and premotor areas; both clustering methods yielded similar subtypes | No subtype differences in age, sex or any PANSS item scores; Subtype 1 had lower age of onset and lower antipsychotic dose | No |
| Dwyer et al. 2023^18^ | 996 [572 patients and 424 HC]; ~67% male; ~ Avg Age: 26 yrs | FEP | MUSE-derived grey and white matter regional volumes | Applied pretrained HYDRA model from Chand *et al*. 2020)^19^ | 4 | Half the FEP individuals were assigned to the previously defined subtype 1 (32%) and subtype 2 (21%); a small percentage (9%) of FEP individuals were assigned to subtype 3 which had mixed subtype 1 and 2 features and the remainder were not assigned to any subtype | No subtypes differences in age and sex; subtype 1 had lower PANSS scores and included greater proportion of individuals prescribed typical antipsychotics | No |
| Jiang et al. 2023^20^ | 2170 [1124 patients and 1046]; ~55% male; ~ Avg Age: 32 yrs | Mixed: Established and FEP | AAL-derived GMV regional measures | SuStaIn | 2 | Both subtypes involved spatially overlapping GMV loss across cortical and subcortical regions; In subtype 1; early GMV reductions were mostly prefrontal and in subtype 2 mostly subcortical | Across both subtypes PANSS negative scores were higher with advancing gray matter loss and the opposite was the case with PANSS positive symptom scores | Yes |
| Shi et al. 2023^22^ | 673 [355 patients and 318 HC]; ~52% male; ~ Avg Age: 27 yrs | Early Stage | Normative W-scores for Brainnetome atlas derived regions following TBM | Sparse K-means clustering | 2 | Both subtypes showed widespread TBM value reduction in cortical and subcortical areas; Subtypes differed in the degree of reduction with subtype 2 having more extensive subcortical reductions; Cerebellar TBM values were higher subtype 1 and lower in subtype 2 | No subtypes differences in age, sex, illness duration and antipsychotic dose; Subtype 2 had a higher PANSS negative symptom and total score | Yes |
| Yao et al. 2023^28^ | 206 [100 patients and 106 HC]; ~66% male; ~ Avg Age: 14 yrs | Childhood and adolescent onset psychosis | Morphometric similarity networks (MSN) derived from measures of surface area, cortical thickness, gray matter volume, Gaussian curvature, and mean curvature | HYDRA | 2 | Subtype 1: reduction in global MSN strength and in frontoteporal and insula regions couple with decreased MSN strength in ventral prefrontal and occipital regions;  Subtype 2: normal global MSN strength, increased MSN strength in the superior frontal cortex and decrease MSN in the paracentral cortex | No subtype differences in age, sex, PANSS subscale scores | No |
| Jiang et al. 2024^11^ | 11250 [4222 patients and 7038 HC]; ~55% male; ~Avg Age: 33 yrs | Mixed: FEP but mostly Established | AAL-derived GMV regional measures | SuStaIn | 2 | Both subtypes showed reductions in cortical thickness and volume and subcortical volume; the magnitude of cortical reductions was greater in frontal regions for subtype 1 and in temporal regions for subtype 2; the magnitude of subcortical reductions was greater in subtype 2 than in subtype 1 except for the striatum which was enlarged in subtype 1 and reduced in subtype 2 | No subtype differences in difference in age, sex, illness duration or any of the PANSS subscale scores | Yes |
| Sone et al. 2024^32^ | 250 [177 patients and 73 HC]; ~59% male; ~Avg Age:42 yrs | Established | Regional FreeSurfer-derived measures of cortical thickness and subcortical volume | SuStaIn | 3 | Subtype 1: Subcortical volume reduction;  Subtype 2: widespread reduction in cortical thickness and increase in GP volume;  Subtype 3: widespread reduction in cortical thickness | No subtype differences in PANSS scores or proportion of treatment resistant cases or proportion of patients prescribed clozapine | No |
| Yakimov et al. 2024^23^ | 239 [132 patients and 107 HC];  ~71% male; ~Avg Age:37 yrs | Established | FreeSurfer-derived measures of the choroid plexus, lateral and third ventricles, regional subcortical volumes, brainstem volume and selected measures of cortical volume | K-means Clustering | 3 | Subtype 1: increased choroid plexus and ventricular volumes;  Subtype 2: lower choroid plexus and ventricular volumes;  Subtype 3: moderately increased choroid plexus and lower ventricular volumes | No subtype differences in age, sex, duration of untreated psychosis or duration of illness, PANSS subscale scores, proportion of treatment resistant cases, body mass index and antipsychotic dose; all subtypes showed cognitive impairment which was greater for subtype 1 | No |
| **Diffusion Tensor Imaging** | | | | | | | | |
| Sun et al. 2015^14^ | 223 [113 patients and 110 HC]; ~51% male; ~Avg Age: 23yrs | FEP | FA and MD of white matter tracks | Agglomerative hierarchical clustering | 2 | Subtype 1: Widespread reduction in FA coupled with increased MD in nearly all tracts;  Subtype 2: Mostly localized FA reduction in the SLF and MD increase in the CST | No subtype differences in difference in age, sex, illness duration or the PANSS positive subscale scores; subtype 1 had higher PANSS negative subscale score | No |
| ^1^ The sample size reported is that of the primary/discovery sample; numbers reflect the sample included in the analyses, which may be smaller than the original sample following quality control or other exclusions; ^2^The clinical stage of patients is not operationally defined across studies; accordingly the term First Episode Psychosis is used when the sample is described as such in the original study, the term Early Stage is used for patients with duration of illness less than 5 years as inferred from the sample demographics; the term Established Stage is used for samples with longer illness duration; ^3^Reductions or increased are typically referenced to the healthy control group; ^4^The associated features reported are those examined in each original study; Avg: average; CHIMERA: Clustering of heterogeneous disease effects via distribution matching of imaging patterns; CSF: Cerebrospinal Fluid; CST: Corticospinal Tract; DTI: Diffusion Tensor Imaging; FA: Fractional anisotropy; FEP: First Episode Psychosis; GAF: Global Assessment of Function; GP: Globus Pallidus; GMV: Gray Matter Volume; HC: Healthy Controls; HYDRA: Heterogeneity through discriminative analysis; ICA: Independent Component Analysis; IDSCN: Individual Differential Structural Covariance Network; MAGIC: Multi-scale semi-supervised clustering; MD: Mean Diffusivity; MUSE: Multi-atlas region Segmentation utilizing Ensembles; PANSS: Positive and Negative Syndrome Scale; PCA: Principal Component Analysis; SLF: Superior Longitudinal Fasciculus; SBM: Source Based Morphometry; SuStaIn: Subtype and Stage Inference; TBM: Tensor Based Morphometry; VBM: Voxel based Morphometry; yrs=years | | | | | | | | |
