## Supplementary Material for "Subtyping Schizophrenia Using Brain Imaging: A Critical Appraisal of Clustering-Based Models"

**Literature Search and Study Selection**

We conducted a structured review of original, peer-reviewed studies published between January 1, 2015, and December 31, 2024, that applied clustering techniques to neuroimaging data in individuals with schizophrenia or related spectrum disorders. Our primary focus was on unsupervised clustering applied to neuroimaging features, alone or in combination with cognitive, clinical, or genetic data, to identify biologically or phenotypically meaningful subtypes at the individual level using the following query:

(schizophrenia) AND (subtype OR cluster) AND (subtypes OR subgroups) AND (neuroimaging OR MRI) AND (english[Language]) AND ("2015/01/01"[Date - Publication] : "2024/12/31"[Date - Publication]) AND (full text[sb]) NOT (review[Publication Type] OR meta-analysis[Publication Type] OR "systematic review"[Publication Type] OR editorial[Publication Type] OR comment[Publication Type] OR letter[Publication Type] OR preprint[Publication Type] OR bipolar[Title] OR "cognitive subtypes")

This query was then adapted for other databases (Embase and Google Scholar) by modifying the search syntax to align with each platform’s controlled vocabulary (and indexing structure, while retaining the core logic and topical focus. This search was supplemented using Consensus.ai, an AI-powered scientific search platform that retrieves literature based on semantic relevance and claim-level indexing.

The PRISMA Flowchart is shown below. Explanatory notes are as follows

1 Studies excluded based on their abstract due to a variety of reasons including: post mortem studies (n=6); group-level pattern identification (n=1); classification (schizophrenia vs controls) (n=3); included patients with psychiatric disorders other than schizophrenia (n=9); included clinical high-risk individuals only (n=3); other (e.g. methods development, radioligand validation; studies examining risk factors for psychosis (n=38). Total number of studies excluded at this stage: 60
2 Studies were excluded because they involved: predefined subgroups of schizophrenia patients based on their clinical (e.g., treatment resistant vs treatment non-resistant), cognitive or other features (n=64); data driven clustering of patients with schizophrenia using non-imaging data (e.g., symptom scores or cognitive task performance) (n=40); clustering was applied to samples that jointly included patients with schizophrenia and other diagnostic labels (n=14); input features included non-imaging data (n=3). Total number of studies excluded at this stage: 121

A full list of all excluded papers is available upon request.

**Identification of studies via databases**

Records removed *before screening*:

Records removed for other reasons (n =0)

Records identified from:

Databases (n =199)

**Identification**

Records screened

(n =199)

Records excluded^1^

(n = 60)

Reports sought for retrieval

(n =139)

Reports not retrieved

(n =0)

**Screening**

Reports excluded^2^:

No data-driven subgroups (n=64)

Clustering using non-imaging features (n = 40)

Transdiagnostic samples (n=14)

Clusering using multidomain input data (n = 3)

Reports assessed for eligibility

(n =139)

Studies included in review

(n = 18)

**Included**
